# Self-supervised contrastive learning improves machine learning discrimination of full thickness macular holes from epiretinal membranes in retinal OCT scans

**DOI:** 10.1101/2023.11.14.23298513

**Authors:** Tim Wheeler, Kaitlyn Hunter, Patricia Garcia, Henry Li, Andrew Thomson, Allan Hunter, Courosh Mehanian

## Abstract

There is a growing interest in using computer-assisted models for the detection of macular conditions using optical coherence tomography (OCT) data. As the quantity of clinical scan data of specific conditions is limited, these models are typically developed by fine-tuning a generalized network to classify specific macular conditions of interest. Full thickness macular holes (FTMH) present a condition requiring timely surgical intervention to prevent permanent vision loss. Other works on automated FTMH classification have tended to use supervised ImageNet pre-trained networks with good results but leave room for improvement. In this paper, we develop a model for FTMH classification using OCT slices around the central foveal region to pre-train a naïve network using contrastive self-supervised learning. We found that self-supervised pre-trained networks outperform ImageNet pre-trained networks despite a small training set size (284 eyes total, 51 FTMH+ eyes, 3 slices from each eye). 3D spatial contrast pre-training yields a model with an F1-score of 1.0 on holdout data (50 eyes total, 10 FTMH+), compared ImageNet pre-trained models, respectively. These results demonstrate that even limited data may be applied toward self-supervised pre-training to substantially improve performance for FTMH classification, indicating applicability toward other OCT-based problems.

**Author Summary:** Full thickness macular holes (FTMH) are a sight-threatening condition that involves the fovea, the area of the eye involved in central vision. Timely diagnosis is paramount because of the risk of permanent vision loss. In clinical practice, full thickness macular holes are commonly diagnosed with the aid of optical coherence tomography (OCT) images of the fovea. However, certain conditions such as pseudoholes and epiretinal membranes may complicate the diagnosis of full thickness macular holes on imaging. Here, we employ the use of artificial intelligence and present a machine-learning model for full thickness macular hole classification and distinction from conditions that may present similarly upon image review. Despite training our model with a smaller data set, it outperformed traditional models previously seen in other works. We provide a strong framework for a self-supervised pre-trained model that can accurately distinguish full thickness macular holes from epiretinal membranes and pseudoholes. Overall, our study provides evidence of the benefit and efficacy with the introduction of artificial intelligence for image classification.

## Introduction

Optical coherence tomography (OCT) allows detailed anatomical imaging via high resolution scans through several hundred microns of retinal tissue layers. The macula is a 5.5 mm wide area of the posterior retina and includes a smaller 1.58 mm wide area known as the fovea which correlates with the eye’s central and highest acuity vision. OCT scanning of the macula generates a wealth of data on possible pathologies present within the retina, but to extract clinically relevant information requires subspecialty review. This imposes an increased workload on ophthalmologists, taking time away from higher level decision-making and patient care.

One condition of interest is full thickness macular holes (FTMH), wherein all layers of the neurosensory retina (between the internal limiting membrane and the retinal pigment epithelium (RPE)) in the central macula (*i.e.*, the fovea) are disrupted. Due to involvement of the fovea, this results in impairment of central visual acuity [1].

The resultant reduced vision worsens with increasing delay in diagnosis and can portend permanent damage the longer surgical repair is postponed compared to earlier interventions [2]. A confounding comorbidity to FTMH is epiretinal membrane (ERM), in which a fibrotic contractile plaque grows over the retina that may reduce anatomic changes similar in appearance to FTMH (*e.g.*, pseudoholes or lamellar holes). ERMs are sometimes associated with macular edema which distorts retinal anatomy further, complicating FTMH diagnosis.

The need for accurate and timely diagnosis of FTMH can be critical for restoring vision, prompting a need for timely assessment of OCT scans. Deep learning has emerged as a promising avenue of research for creating automated expert reader models to assess medical scans in-line with the acquisition process. Briefly, deep learning refers to (artificial) neural networks that are many layers deep [3]. Neural networks, in turn, are defined as layers of nonlinear signal processing units (artificial neurons) connected by weights (artificial synapses). The effect of training a neural network is to adjust the weights such that the system approximates a function mapping a given input to a desired output. A multi-layered network of nonlinear processing units can in theory approximate any function to arbitrary precision with sufficient training data [4]. Convolutional neural networks (CNN) are particularly well-suited to image recognition tasks because the shared-weight architecture of convolutional kernels learns translation-covariant visual features automatically [5].

Developing a functional deep learning model requires large quantities of well-annotated ground truth data, and a training regimen that allows the model to learn discriminative features for the problem at hand. A frequent issue arising in training deep learning models with medical data is low dataset size. This makes it impractical to train a naïve network from scratch and has typically been circumvented by fine-tuning a pre-trained network (transfer learning). This can also be problematic, as pre-training is usually based on images from a non-medical domain, typically ImageNet, which means that features extracted by the network may not be relevant to the specific nuances of the tissue and imaging method. An alternative to transfer learning for handling low data volume is self-supervised learning, which learns a suitable image representation by performing a pretext task on unlabeled data, which is usually available in larger quantities than labeled data. Various approaches to self-supervised learning have been introduced, many involving an encoder, which is a feedforward neural network that represents an image by a vector (also known as an embedding). Contrastive learning pushes positive pairs closer and negative pairs farther apart in the embedding space. A primary exemplar of this contrastive learning approach is SimCLR [6, 7].

The last several years have seen an explosion of deep learning models applied to ophthalmic clinical technologies including OCT and fundus imaging. These applications may be divided into the broad areas of classification/diagnosis [8–18], segmentation [19–26], image quality [27], and demographics prediction [28]. The current ophthalmic deep learning models focus primarily on diabetic retinopathy, age-related macular degeneration, retinopathy of prematurity, and glaucoma [29–31].

Deep learning based OCT image analysis for FTMH has also received attention lately, with models for classification [16, 17], segmentation [23–26], and prognosis of success for FTMH corrective surgery [18, 32, 33]. A review is also available [34]. Owing to the paucity of labeled FTMH data, the majority of these models are pre-trained on ImageNet, and subsequently fine-tuned on a small amount of labeled FTMH OCT images. This transfer learning scheme was adopted in developing the classification model by Pace et al. [16], which achieves 95% accuracy in distinguishing between normal, Drusen, and FTMH images. Carvalho et al. [17], demonstrated an accuracy of 90.6% for FTMH identification, although modeling specifics are not provided. While these results are good, there is room for improvement.

We employ a pre-training method based on a variant of SimCLR that leverages 3D information, and which is tailored to small datasets with multi-slice imaging modalities such as OCT, described here as Random Slice Contrastive Learning (RaSCL). Our model achieves robust feature recognition in OCT scans for assessing FTMH that can outperform traditional ImageNet pre-trained models.

## Materials and Methods

### Patient data

Data used in this study was acquired from patients prior to surgical intervention [35]. Patients with a history of ocular trauma, amblyopia, recent ocular surgery (within three months), severe ocular myopia, any diabetic macular retinopathy, and retinal pathology associated with systemic conditions (uncontrolled hypertension) were excluded.

The dataset contains OCT images of 61 eyes from 60 patients with FTMH (46 Females, 15 Males, ages 52-84 years, mean: 69.6 years, SD: 6.4 years) with confirmation of diagnosis and image quality assessed by three trained readers. The remainder of FTMH-negative control data consisted of scans from patients diagnosed with ERM, comprising 274 eyes from 264 patients (140 female, 134 male, ages 23-93 years, mean: 70.5 years, SD: 8.6 years).

A B-slice is a horizontal linear OCT scan, producing a 2D image of a macular tissue section. Typically, multiple uniformly spaced B-scans are acquired, with the central slice going through the fovea. B slices were individually labeled as having FTMH by consensus of three expert readers. B-slices were exported from Heidelberg Spectralis OCT instruments (Heidelberg, Germany) as TIFF files at 496 × 512 resolution, with pixel size 3.87 × 11.38 µm at the retina. Macular OCT scan protocols available included B-scan slices at either 243 *μm* or 121 *μm* apart, and both protocol types were included.

#### Test splits

Test sets for model evaluation were assembled from 15% of total eyes, comprising 10 FTMH and 40 control eyes, using only the central B-slice to ensure consistency between eyes with differing numbers of B-scans that show evidence of FTMH. Three random split replicates with disjoint test sets were generated for statistical validation. Within each replicate, self-supervised pre-training and fine-tuning were performed using only the training split. For each replicate, training sets were further divided into 8-fold cross-validation splits.

#### Data stratification

Data was stratified based primarily on diagnosis (FTMH or ERM). Secondary features of age, sex, and pre-operative vision were used to stratify data by optimizing the mean and standard deviation of each subset relative to the overall dataset mean and standard deviation for each feature (treating sex as ordinal).

### Preprocessing

B-slices were cropped and resized to 224 × 224 resolution, and then augmented with random noise, brightness, contrast, cropping, and horizontal flips. Additive noise was generated by sampling a normal distribution with randomly drawn parameters µ ∈ [−0.1,0.15] and σ^2^ ∈ [0,0.2], cropping values to the interval [0,1], to mimic noise patterns typical of B-slices. Random crops comprised 50-100% of the original scan area.

### Architecture

The model architecture consists of a CNN followed by a multi-layered perceptron (MLP) based on the SimCLR approach [6, 7]. The CNN uses a naïve ResNet50 framework at standard width [36]. The MLP comprises three fully connected layers of 512 nodes each. The final layer is used as a projection head during pre-training, which is fed to a classifier head during fine-tuning. Models were trained for 800 epochs during pre-training and 500 epochs during fine-tuning and were validated every 10 epochs. An 8-fold training set split was used for cross-validation training and unweighted ensemble averaging.

Training was performed on a workstation with an NVIDIA RTX A6000 GPU, a 24-core Intel Xeon W 3345 CPU, under Ubuntu 20.04, using Python 3.8.10 and TensorFlow 2.9.0. Taken together, pre-training and fine-tuning ran for 7.5 hours per ensemble replicate in this computational framework.

### Self-supervised pre-training

Pre-training was performed using contrastive self-supervised learning. Image slices from the same eye constituted positive pairs, while images from unrelated eyes constituted negative pairs. In more detail, for each batch, a B-slice was randomly selected from an eye, then a neighboring slice at a distance up to 2 slices away on either side was randomly selected (uniformly) as a positive pair. Scans between unrelated eyes constituted negative pairs. We refer to our scheme as 3D spatial contrast or RaSCL, which is similar in spirit to the SimCLR adaptation in Gomariz, *et al.* [26]. However, in the latter, positive pair slice distance is distributed normally: *d*∼𝒩(0, 0.25*μm*), and since their B-slices are separated by 111*μm*, their scheme is effectively the same as standard SimCLR.

The model transferred for fine-tuning was selected based on the training epoch with maximal accuracy and minimal loss, *i.e.*, min(loss/accuracy), on the validation set. Pre-training data included three B-slices per eye centered on the fovea.

### Supervised fine-tuning

Fine-tuning was performed using 8-fold cross-validation. Splits were stratified by diagnosis, age, sex, and visual acuity. CNN weights were fixed during fine-tuning, with only the MLP tuned using a binary cross-entropy loss function, with the two classes specified as control and FTMH. The best model per split was selected by the validation epoch with minimum validation loss divided by validation accuracy. The 8-fold split model outputs were combined as an averaged ensemble with equal weights between components. Fine-tuning data included up to three B-slices per eye centered on the fovea.

## Results

Ensemble models were evaluated on the holdout test set comprising 10 FTMH B-slices and 40 control B-slices, performed in triplicate for different train-test replicates (Fig. 1).

**Fig. 1.**
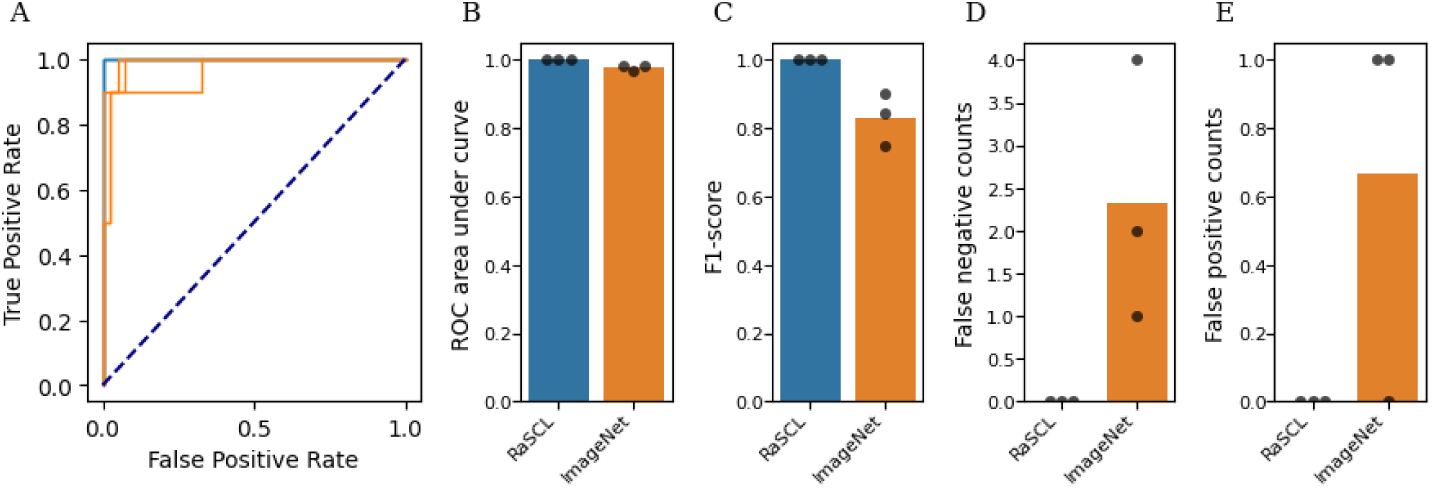
Summary statistics on three holdout replicates evaluated by models pre-trained with RaSCL or ImageNet. (A) Receiver operating characteristic (ROC) curves with associated area under curve (B). Overall F1-score (C). False negative (controls, out of 40) and false positive (FTMH, out of 10) counts (D, E). Bars indicate mean statistics for each group, and dots indicate individual replicates.

### Performance

The 3D spatial contrast pre-trained network performed completely accurately, diagnosing all test scans correctly in all replicates. The ImageNet pre-trained model performed significantly worse, in terms of both accuracy (*p* = 0.017) and F1-score (*p* = 0.018). Test samples were classified as FTMH or control using a default softmax score threshold of 0.5.

### Gradient visualization

Visualization of gradient activation for an FTMH input (Fig. 2A) shows that for the RaSCL model, strong activation is localized around the macular hole (Fig. 2A’). In contrast, the ImageNet model presents a weaker saliency around the middle of the B-slice (Fig. 2A’’). Gradient activation for a control input (Fig. 2B) shows that the RaSCL model highlights the inner limiting membrane on the upper surface of the macula, and the undisrupted retinal pigment epithelium below (Fig. 2B’). ImageNet similarly presents weak saliency for this scan (Fig. 2B’’).

**Fig. 2.**
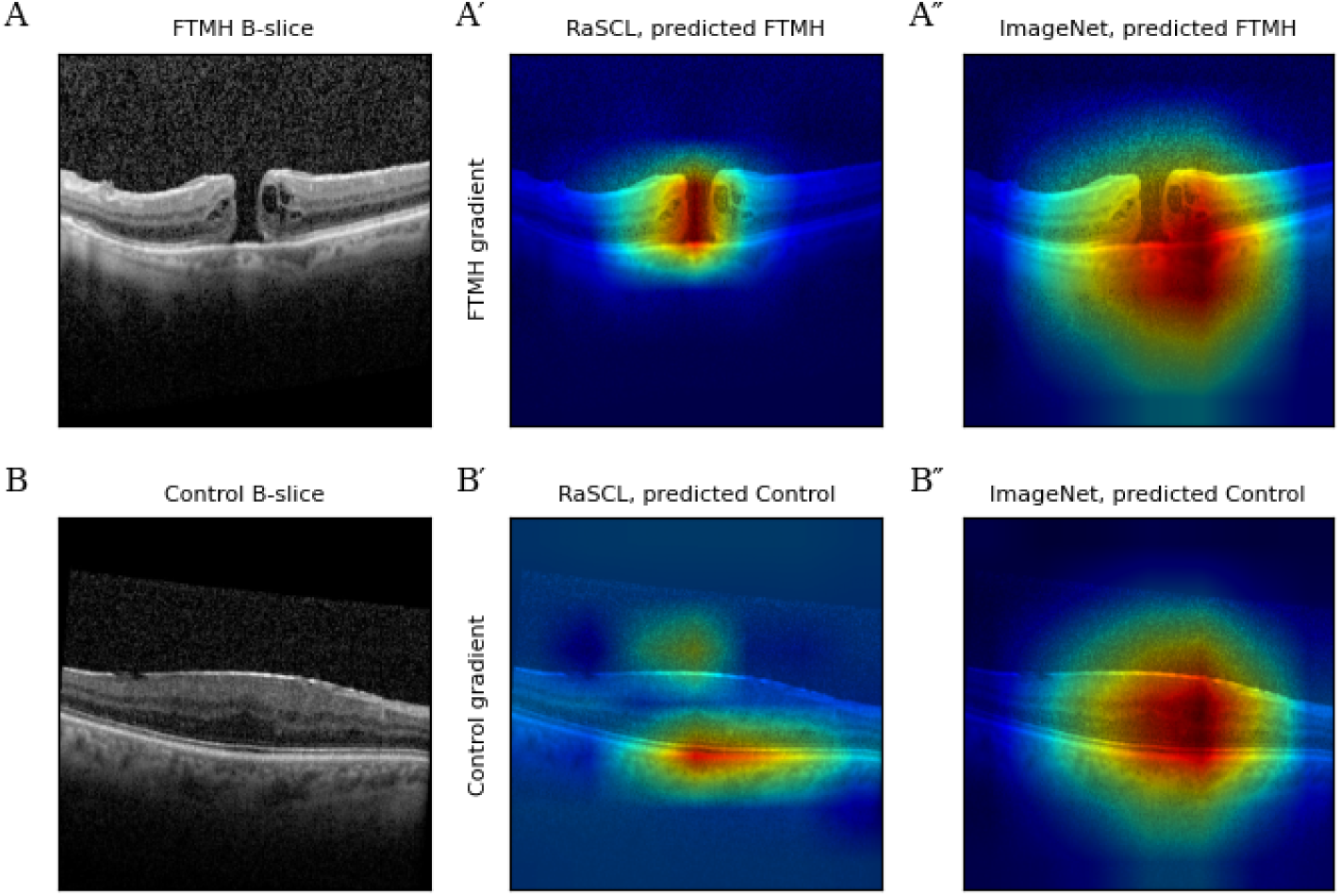
Gradient visualizations of FTMH and control B-slice images for RaSCL and ImageNet pre-trained models. (A) Gradient visualization of the FTMH output unit using an FTMH B-slice. (B) Gradient visualization of the control output unit using a control B-slice.

### Challenge images

A challenge dataset was assembled comprising 34 images presenting lamellar holes, which are partial-thickness defects that can appear similar to FTMH, but do not extend through all the neuronal layers to the RPE. Most lamellar holes do not require intervention. Pseudoholes are another example of a visually similar diagnosis without the need for clinical intervention. We challenged all replicates of each model type and found that all replicates of the RaSCL pre-trained models correctly classified all the challenge data, while the ImageNet pre-trained models misclassified several images, with a mean accuracy of 0.971, *p* = 0.158. We visualized the FTMH gradients to explore possible explanatory factors for the misclassifications (Fig. 3B-D). While RaSCL FTMH gradients had strong activation around the pseudohole (Fig. 3C), this activation did not extend down to the RPE and did not lead to misclassification. By contrast, ImageNet FTMH gradients were diffused around the tissue, suggesting poor saliency from the domain transfer model (Fig. 3D).

**Fig. 3.**
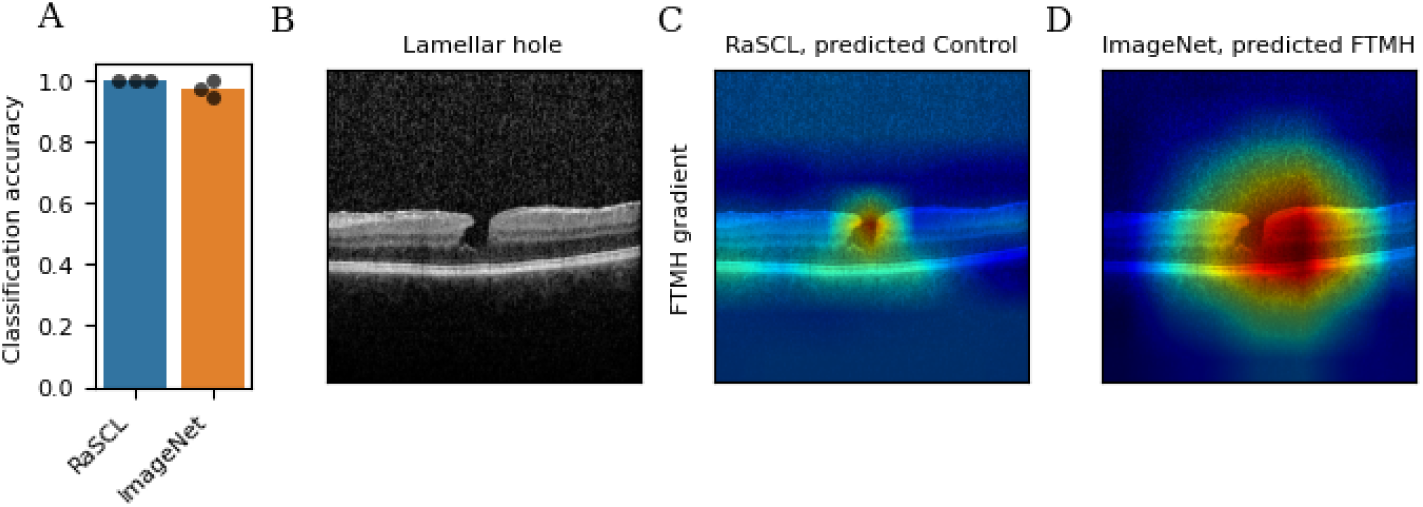
Challenge image set performance. (A) Classification accuracy. Bars indicate mean and dots indicate replicates. (B) Challenge image with a lamellar hole. (C) Image correctly classified by the RaSCL model overlaid with FTMH gradients. (D) Image incorrectly classified by ImageNet pre-trained models with overlaid FTMH gradients.

## Discussion

This work demonstrates a potentially strong diagnostic model for full-thickness macular holes, which are a serious eye care concern due the significant vision impairment resulting from the characteristic defect in the central fovea and subsequent loss of central visual acuity [1]. The vitreous is a clear gel substance which fills the eye’s posterior chamber and has attachments to the macula. Macular holes develop due to vitreous traction at the vitreofoveal interface and are most commonly idiopathic but can result from traumatic injury to the eye or secondary to other ocular pathology [1, 37]. Gass and Johnson were the first to suggest that this phenomenon occurs due to focal shrinking of the vitreous which results in traction on the fovea [1, 37]. In Gass’ original classification [1, 38], the natural progression of macular holes proceeds through stages associated with worsening visual acuity: foveolar detachment in stage 1, formation of a foveal hole with associated subretinal fluid and cystoid macular edema in stage 2, formation of an FTMH in stage 3, followed by an FTMH with a complete posterior vitreous detachment from the vitreofoveal interface in stage 4.

The use of artificial intelligence (AI) in ophthalmology has advanced significantly, owing to the many image-based investigations in the field. Previous attempts on automated FTMH classification have tended to use supervised ImageNet pre-trained networks with good results but which leave room for improvement. The present work demonstrates that near perfect accuracy is achievable for FTMH classification, given a good image representation learned from the data. The CNN models in this study also present a good starting point for fine-tuning other OCT-related diagnostic and prognostic questions. Prior works addressed whether the outcome of FTMH-corrective surgery can be predicted, with only moderate success [18, 32, 33]. A natural step would be to use the RaSCL approach to develop models for accurate surgical prognosis, which could have clinical benefit by helping to avoid unnecessary surgeries. This study highlights spatial contrastive learning as a powerful pre-training approach for medical scans, which can be applied to deep learning models for other imaging modalities to enhance their tractability.

Random-slice spatial contrastive learning allows the network to develop features that are strongly discriminative for the downstream task of recognizing FTMH. This work demonstrates that strong purpose-specific pre-training is viable for small dataset sizes, and that this method improves performance over transfer learning models. Visualization of gradient activation (Fig. 2) shows sharp saliency in the RaSCL model. The lower discriminative capacity and poor saliency of transfer learning models may also result in brittle classifiers with inferior generalization.

Prior to clinical roll-out, further considerations need to be addressed. This data is limited to one retina practice with four retina surgeons, restricted to one device type, and has a narrow set of inclusion criteria. To develop higher confidence in the model for deployment, a larger dataset including numerous providers, other devices (*e.g.,* Heidelberg Spectralis, Zeiss Cirrus, and Topcon), a patient population with a broader demographic profile, and disease-agnostic OCT data would be required.

We can apply the framework presented here to an expanded dataset to address these limitations. We included challenge data showing performance on lamellar holes (Fig. 3) which is a condition that could be conflated with FTMH. Results on the challenge dataset consisting of lamellar/pseudoholes corroborate the robustness of contrastive pre-training to yield improved performance over non-medical domain pre-trained networks.

The findings in this study shows that deep learning algorithms can be used for computer-assisted screening of FTMH in optometry and primary care settings, promoting appropriate and timely referrals to retinal specialists. The capability improves clinical workflow, reduces clinician workload, streamlines referral so that patients get access to specialist care sooner, increasing overall quality of patient care. AI classification significantly reduces referral processing time allowing patients to be scheduled faster enabling efficient triage and care [30, 39]. However, the small size of the datasets in this study justifies only modest conclusions about model performance. The DL model used for this dataset provides a starting point for other clinically relevant image-based diagnoses in ophthalmology and beyond.

## Data Availability

All relevant data are within the manuscript and its Supporting Information files.

